# Predicting adverse outcomes after cardiac surgery using multi-task deep neural networks, clinical features, and electrocardiograms

**DOI:** 10.1101/2024.12.31.24319813

**Authors:** Chin Siang Ong, Raimon Padrós-Valls, Erik Reinertsen, Steven Song, Katherine Young, Thoralf Sundt, Collin M. Stultz, Aaron D. Aguirre

## Abstract

**Background:** Risk stratification models estimate the probabilities of adverse outcomes after cardiac surgical procedures, which helps clinicians and patients make informed decisions.

**Objectives:** We used the 12-lead electrocardiogram (ECG) and/or Society for Thoracic Surgeons (STS) variables to predict postoperative outcomes using deep learning methods that can incorporate diverse data types.

**Methods:** We developed a deep convolutional neural network (“ECGNet”) that predicts operative mortality and other adverse outcomes using preoperative 12-lead ECGs (n=30,877) from 12,933 patients who underwent 13,299 cardiac surgical procedures. We also developed a deep neural network applied to preoperative STS variables (“STSNet”). STSNet and ECGNet are multi-task neural networks that utilize secondary outcomes to augment prediction of mortality using the same neural network.

**Results:** ECGNet achieved a mean area under the receiver operating characteristic curve (AUC) of 0.85 for predicting operative mortality for all procedures and 0.93 for valve procedures. STSNet achieved a mean AUC of 0.85 for all procedures, with statistically similar performance as ECGNet for all procedures. Combining ECGNet and STSNet achieved a mean AUC of 0.90 for predicting operative mortality after all procedures, which is significantly higher than either ECGNet or STSNet alone.

**Conclusions:** A deep neural network trained on STS features has higher predictive performance than previously reported for existing conventional models and is not limited to certain types of cardiac surgical procedures. A model trained on ECG alone can predict operative mortality with similar performance as STS features and adding ECG to STS features in a neural network can improve performance. These findings demonstrate the potential in leveraging deep learning on multidimensional data sources to predict outcomes after cardiac surgery.

**Condensed abstract:** In this study, deep learning (DL) is applied to electrocardiograms and clinical features used in the standard STS risk prediction tools to generate new high-performing risk calculators for cardiac surgical procedures. Preoperative voltage waveforms contain information about cardiovascular risk and cardiac function and are passed as inputs to the deep learning model. These risk models apply to all cardiac procedures including those procedures that do not have standard STS risk calculators and provide improved performance. DL models enable the incorporation of additional modalities of data to improve risk prediction in cardiac surgery.

## Introduction

Outcome prediction models have been developed in cardiac surgery for various purposes including quality measurement, practice improvement, and clinical decision making (1). These models typically utilize laboratory values, indicators of clinical status, and other data from the preoperative period. The Society of Thoracic Surgeons (STS) developed one of the most frequently used risk models to predict outcomes after cardiac surgery, including operative mortality (2).

The electrocardiogram (ECG) provides a rich spatiotemporal representation of electrical activity in the heart. As a routine part of preoperative cardiovascular testing, the ECG may provide prognostic information in patients with active cardiac conditions (3). The ECG contains features related to many risk factors for adverse post-surgical outcomes, including rhythm abnormalities, structural defects, and coronary artery disease (3). The ECG waveform may therefore provide powerful information for preoperative risk prediction. However, to our knowledge, no cardiac surgical outcome risk model uses waveform data from the ECG.

Convolutional deep neural networks (DNNs) efficiently capture the spatial dependencies between pixels in an image via convolution functions. Originally developed for computer vision tasks, convolution DNNs have been adapted and applied to ECGs to interpret rhythms with cardiologist-level accuracy (4), and to detect traditionally non-obvious cardiac conditions from the ECG alone such as reduced ejection fraction (5-7), the likelihood of recent atrial fibrillation (8), or the estimation of central cardiac pressures (9). DNNs learn non-linear combinations of various parts of the input signal that are correlated with the outcome of interest (10).

We hypothesized that the voltage waveform in the preoperative ECG contains prognostic information for postoperative outcomes. To test this hypothesis, we trained a multi-task convolutional DNN, “ECGNet”, to predict operative mortality and other adverse outcomes from preoperative ECGs obtained from patients who underwent cardiac surgical procedures. Additionally, we trained a multi-task DNN “STSNet” using the standard STS features and evaluated the value of adding ECG data to STS features. These multi-task neural networks also use secondary outcomes, including stroke and renal failure, to enhance prediction of mortality.

## Methods

### Data sources and study population

We extracted and digitized the electrical waveform information from 4.7 million (representing the entirety of available data) 10-second 12-lead resting ECGs from the GE MUSE system (Chicago, IL) at the Massachusetts General Hospital (MGH) (Boston, MA). ECGs were accessed from the manufacturer’s raw data format and sampled at either 250 or 500 Hz. To train and evaluate the algorithm on a consistent number of measurements, ECGs sampled at 500 Hz were downsampled to 250 Hz; all ECGs were thus composed of 2500 samples per lead. The final data representation of an ECG was a 2500 x 12 (12 leads with 2500 samples each) matrix. ECGs with zero values for more than 25% of the waveform were considered noisy and discarded. Forty-eight features used in existing STS risk models for operative mortality were extracted from the STS dataset at the MGH (Supplementary Methods, Supplementary Table 1). Continuous features were normalized via subtracting the population mean and dividing by the population interquartile range. ECGs were standardized by subtracting the mean and dividing by the standard deviation from the training set. Each ECG lead was standardized independently. Standardization was performed using the summary statistics of the training set. Categorical variables were one-hot encoded.

### Patient Population

Between July 2002 and September 2019, 20,836 cardiac surgical procedures were performed at the MGH. Patients undergoing procedures involving circulatory arrest (n=1,324), or those under the age of 18 (n=80), were excluded, leaving 19,432 patients. This cohort was cross-referenced against the ECG dataset using a 60-day preoperative window; ECGs within the 60-day preoperative window were unavailable in our record system for 6,499 patients, resulting in 12,933 patients with 13,299 surgeries and 30,877 preoperative ECGs (Figure 1).

**Figure 1:**
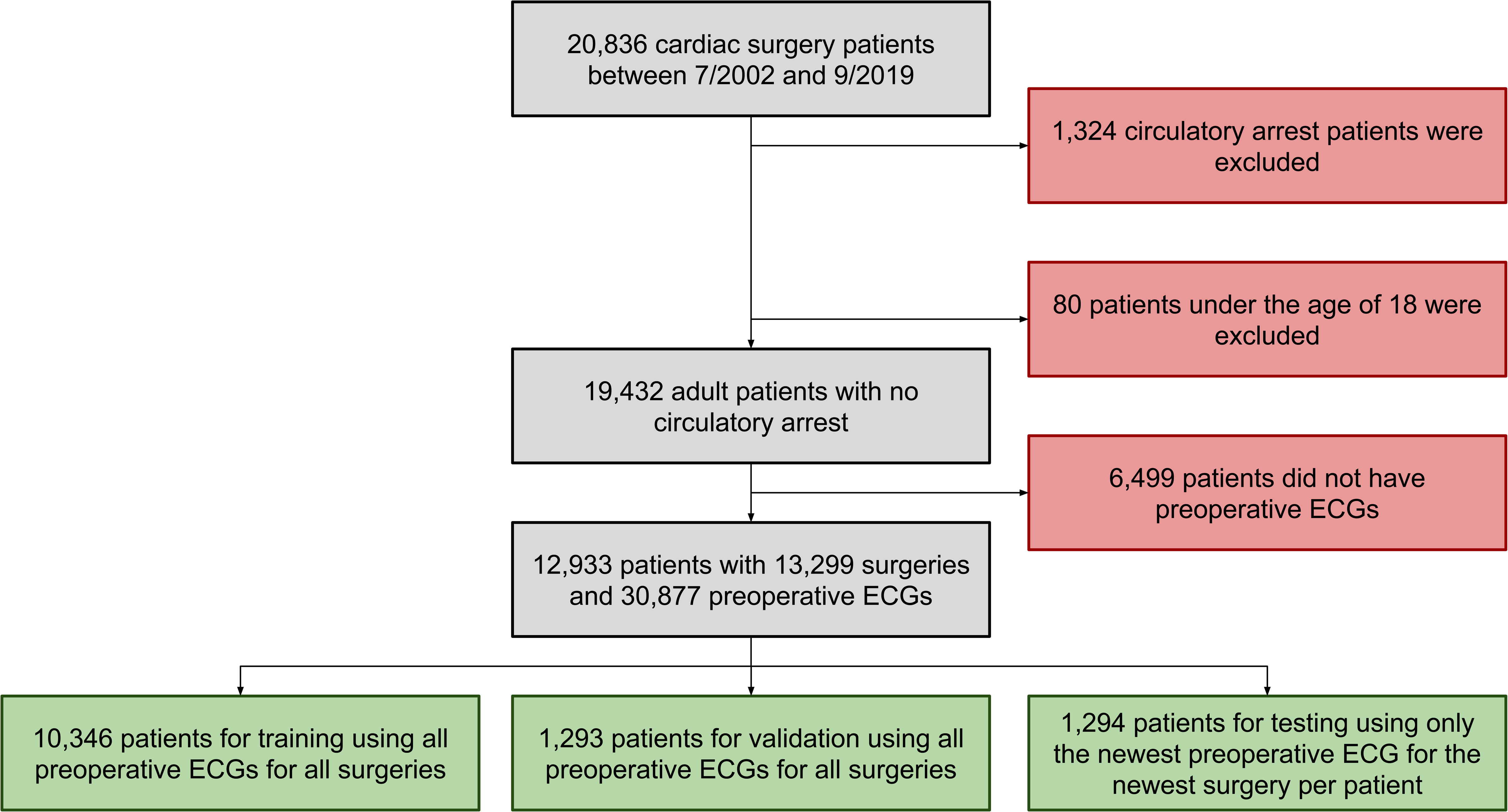
Flow diagram of the inclusion and exclusion criteria applied to the study cohort. Patients who underwent cardiac surgical procedures between July 2002 and September 2019 at the MGH were initially included. Patients who underwent procedures involving circulatory arrest (n=1,324), were under the age of 18 (n=80) or did not have ECGs within the 60-day preoperative window in our record system (n=6,499) were excluded, resulting in 12,933 patients with 13,299 surgeries and 30,877 preoperative ECGs. This final cohort was split into training, validation, and test sets using an 8:1:1 ratio.

### Ethics

This study was approved by the Institutional Review Board of Mass General Brigham.

### Deep learning

Data was processed and loaded via a custom software pipeline written in Bash shell scripts, Python 3.8, and Docker. Deep learning models (Figure 2) were developed and trained using TensorFlow 2.4. We optimized DNN architecture hyperparameters using a random search (Supplementary Methods). Optimal network architectures for tabular (e.g., non-voltage), voltage, or both tabular and voltage input types are described in Supplementary Table 2.

**Figure 2:**
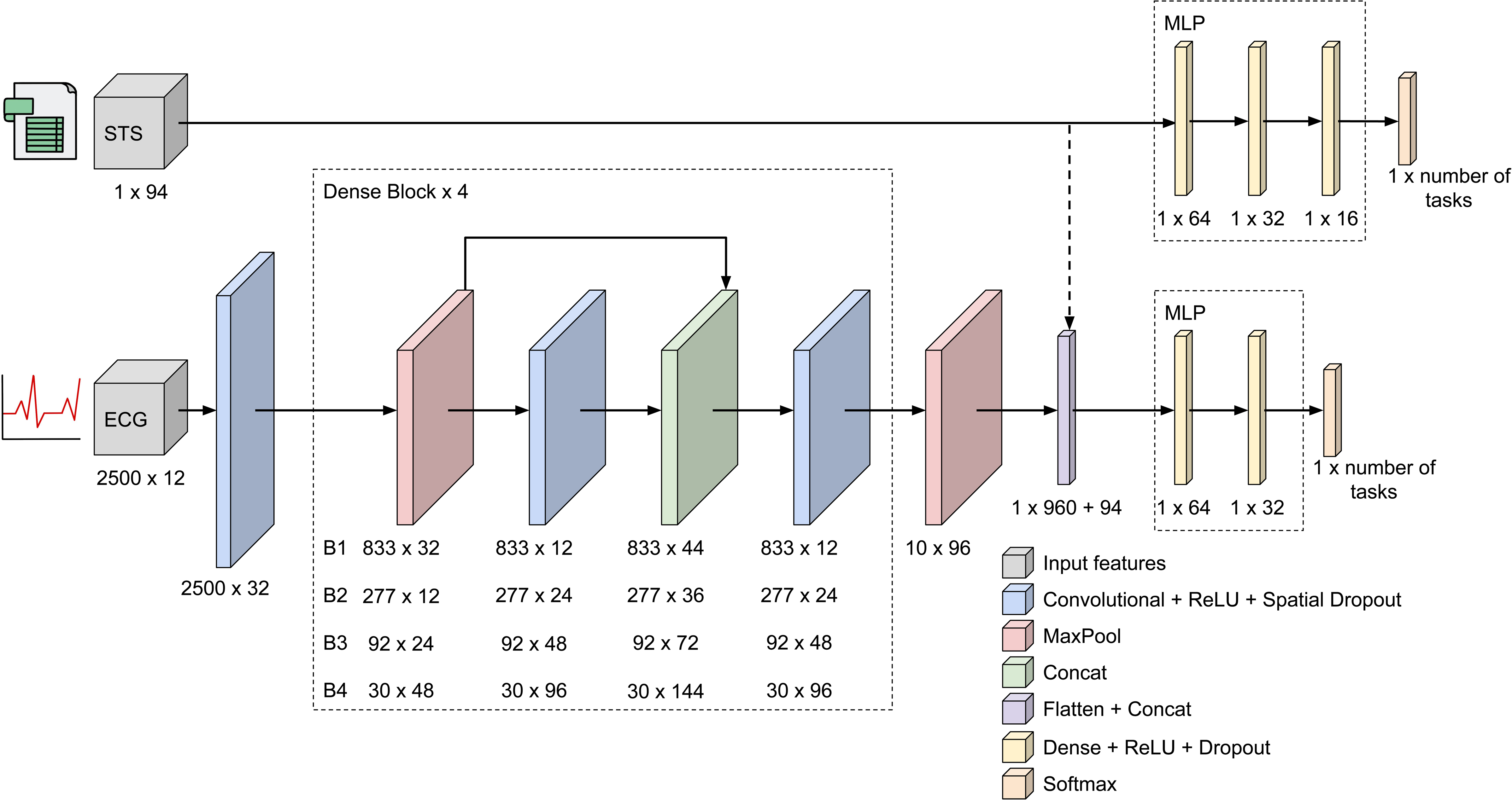
Deep neural network architectures determined via random hyperparameter search. STSNet feeds the STS features in a two-layer DNN (top solid line). In ECGNet, the ECG is processed with a thirteen-layer convolutional block plus a two-layer DNN (bottom solid line). When combining both input data types, the ECG was first processed with a convolutional DNN block, then the STS features were concatenated with the output of this block (dashed line), and finally, the resulting array was processed with a two-layer DNN. Note that the final two-layer DNN for ECGNet is the same as the one when both input data types are combined but different than the one obtained for STSNet.

Models were trained for a maximum of 500 epochs with a batch size of 256. The categorical cross entropy function was used for the loss function. For single-task models, the loss of the outcome of interest was monitored to perform learning rate reduction and early stopping. For multi-task models, the sum of losses of each outcome was monitored to perform learning rate reduction and early stopping. After 16 consecutive epochs without improvement in validation loss, the learning rate was reduced by a factor of two. In models using the AdamW optimizer with weight decay, weight decay was reduced by the same factor and at the same time as the learning rate. Early stopping was performed whereby training concluded after 32 consecutive epochs without improvement in validation loss.

For single-task models, training, validation, and test sets were stratified by the prevalence of the specific outcome of interest, patient age quartiles, and patient gender. For multi-task models, each was stratified by mortality, patient age quartiles, and patient gender. Stratification ensured similar outcome, patient age quartile, and patient gender prevalence across splits. Additionally, data were split amongst training, validation, and test sets on a per-patient, rather per-ECG, basis. This procedure mitigates data leak whereby the same patient contributes ECGs to more than one of the training, validation, or test sets, which can result in models that achieve high performance on the test set due to learning patient-specific features rather than features associated with the label of interest. 80% (n=10,346) of unique patient IDs were allocated to the training set, 10% (n=1,293) to the validation set, and 10% (n=1,294) to the test set. In the training and validation sets, each patient could potentially contribute multiple ECGs if available in the preoperative window; this approach can be considered as a form of data augmentation whereby the model is trained on a variety of similar yet slightly different representations of data from each patient and their corresponding label. Data augmentation can improve generalization of models to out-of-sample data (11). In the test set, only the most recent preoperative ECG was used for each patient, consistent with a potential use case. To obtain more reliable performance metrics, ten different splits (bootstraps) of random training, validation, and test splits were generated, drawing samples without replacement.

Both single-task and multi-task convolutional DNNs and DNNs were trained. When trained in a multi-task setting, the output of each model included predicted probabilities for each of six outcomes: operative mortality, stroke, renal failure, prolonged ventilation, reoperation, and long length of stay (Supplementary Table 3) (1). The output of each model included the predicted probabilities for the specific outcome of interest.

### Model evaluation and statistical methods

The model which achieved the lowest validation loss during training was evaluated on the test set of the bootstrap the model was trained on (1,294 patients). The probability of operative mortality predicted by the model was saved for each patient in the test set. Model performance metrics and all statistical tests were calculated using Python, SciPy and Scikit-learn. Calculations which required predicted events rather than predicted probabilities were performed by binarizing the predicted probabilities of operative mortality around a threshold probability selected via maximization of the F1 score (12). By maximizing the F1 score, precision and recall are balanced equally. The mean and standard deviation of each metric was calculated across ten bootstraps per model. Receiver operator characteristic (ROC), precision-recall (PR), and calibration curves were plotted (Figure 3) when predicting mortality in a multi-task fashion. All models were independently calibrated using an isotonic regressor trained on the validation set. Calibration curves were created by binning the predicted probabilities into five equally sized bins (i.e., pentile where each bin has the same number of patients) within each bootstrap. Calibration slopes and calibration intercepts were calculated by fitting a line to the actual probability of operative mortality within each decile of predicted probability of operative mortality. Statistical comparison of metrics of model performance were done using paired t-tests.

**Figure 3:**
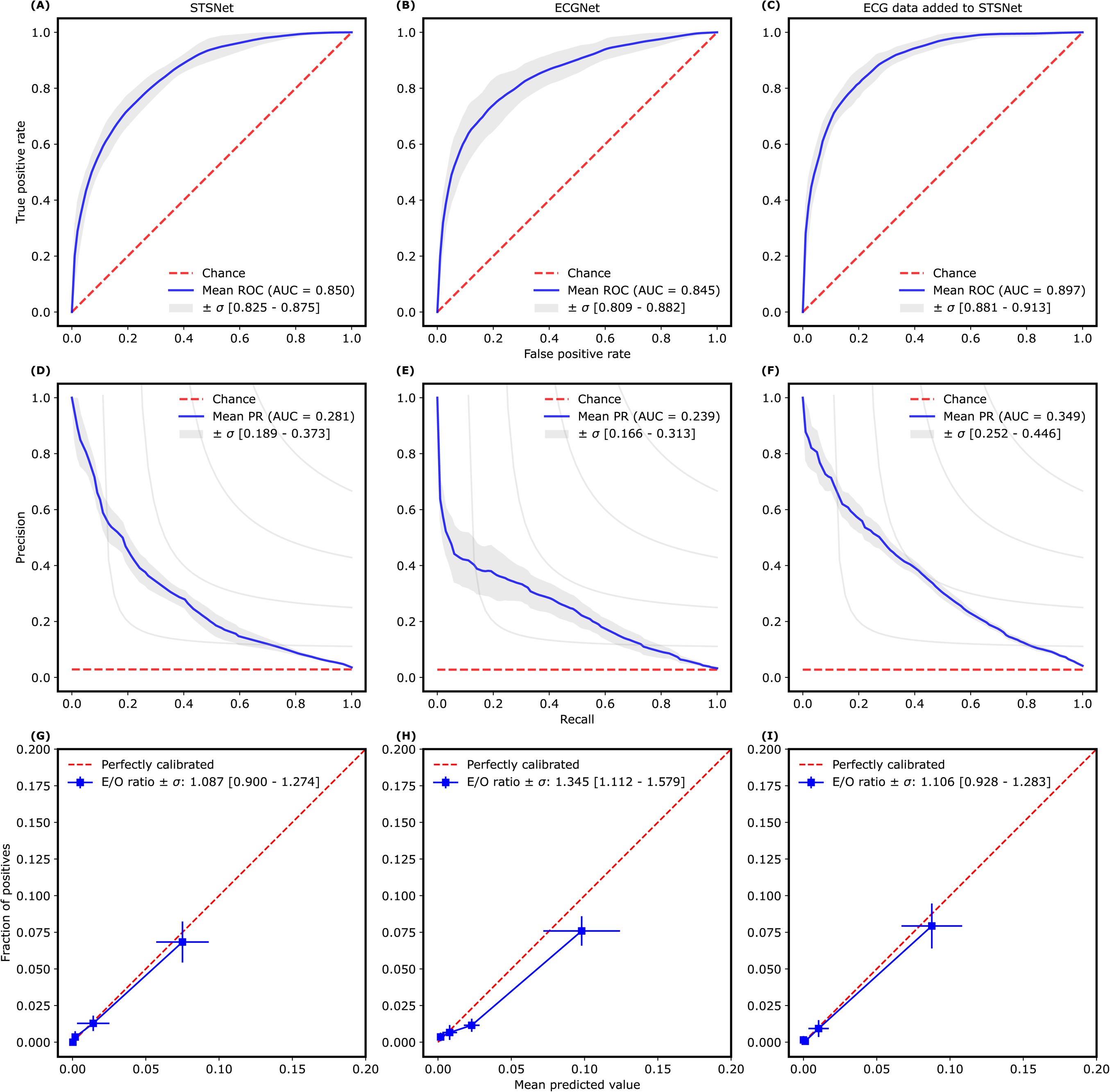
Receiver operator characteristic (ROC), Precision-Recall (PR) and calibration curves. ROC, PR, and calibration curves across ten bootstraps for multitask models predicting operative mortality using (left to right) STS features (STSNet), ECG voltage (ECGNet), and STS features + ECG voltage. Top: ROC curves with mean AUCs ± 1 standard deviation. Middle: PR curves with mean AUPRCs ± 1 standard deviation. Bottom: Calibration curves with mean E/O ratio ± 1 standard deviation.

## Results

Of the 12,933 patients who met inclusion criteria and had preoperative ECGs, 394 (3%) had operative mortality. The differences in patient characteristics between those who died and those who did not are presented in Table 1.

**Table 1:**
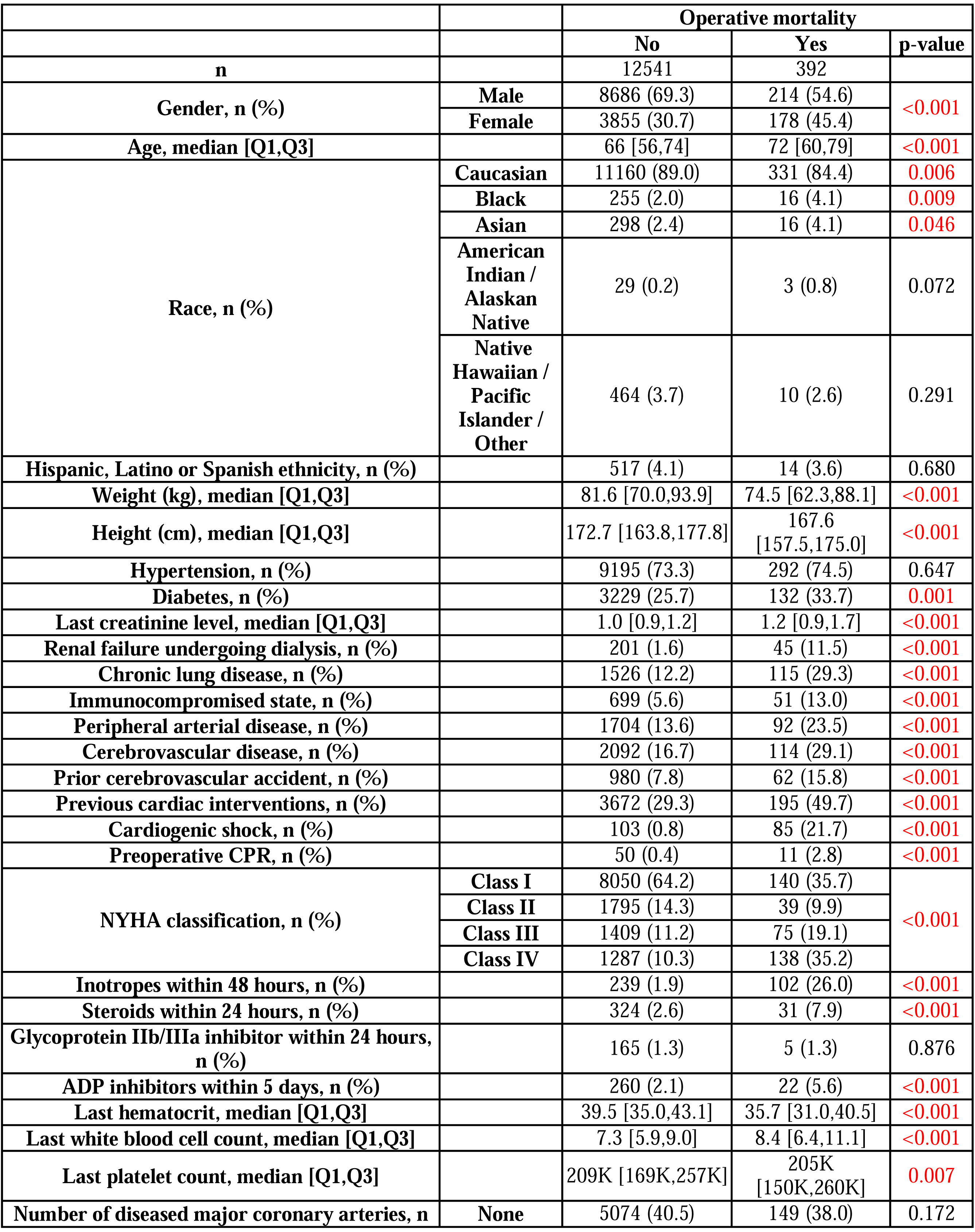
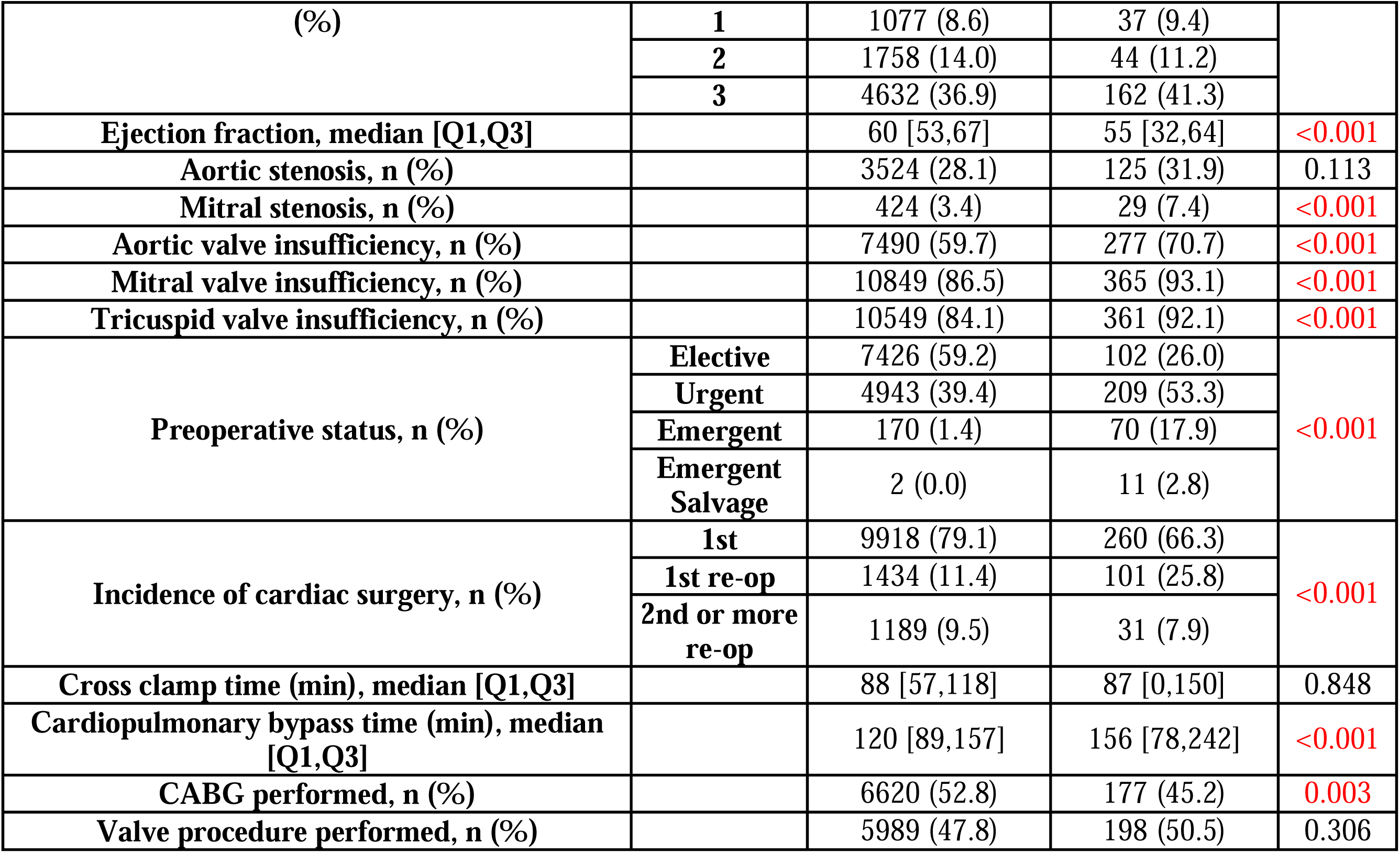
Patient characteristics of the MGH cohort (n = 12,933)

|  |  | Operative mortality |  |  |
| --- | --- | --- | --- | --- |
|  |  | No | Yes | p-value |
| **n** |  | 12541 | 392 |  |
| **Gender, n (%)** | **Male** | 8686 (69.3) | 214 (54.6) | <0.001 |
|  | **Female** | 3855 (30.7) | 178 (45.4) |  |
| **Age, median [Q1,Q3]** |  | 66 [56,74] | 72 [60,79] | <0.001 |
| **Race, n (%)** | **Caucasian** | 11160 (89.0) | 331 (84.4) | 0.006 |
|  | **Black** | 255 (2.0) | 16 (4.1) | 0.009 |
|  | **Asian** | 298 (2.4) | 16 (4.1) | 0.046 |
|  | **American Indian / Alaskan Native** | 29 (0.2) | 3 (0.8) | 0.072 |
|  | **Native Hawaiian / Pacific Islander / Other** | 464 (3.7) | 10 (2.6) | 0.291 |
| **Hispanic, Latino or Spanish ethnicity, n (%)** |  | 517 (4.1) | 14 (3.6) | 0.680 |
| **Weight (kg), median [Q1,Q3]** |  | 81.6 [70.0,93.9] | 74.5 [62.3,88.1] | <0.001 |
| **Height (cm), median [Q1,Q3]** |  | 172.7 [163.8,177.8] | 167.6 [157.5,175.0] | <0.001 |
| **Hypertension, n (%)** |  | 9195 (73.3) | 292 (74.5) | 0.647 |
| **Diabetes, n (%)** |  | 3229 (25.7) | 132 (33.7) | 0.001 |
| **Last creatinine level, median [Q1,Q3]** |  | 1.0 [0.9,1.2] | 1.2 [0.9,1.7] | <0.001 |
| **Renal failure undergoing dialysis, n (%)** |  | 201 (1.6) | 45 (11.5) | <0.001 |
| **Chronic lung disease, n (%)** |  | 1526 (12.2) | 115 (29.3) | <0.001 |
| **Immunocompromised state, n (%)** |  | 699 (5.6) | 51 (13.0) | <0.001 |
| **Peripheral arterial disease, n (%)** |  | 1704 (13.6) | 92 (23.5) | <0.001 |
| **Cerebrovascular disease, n (%)** |  | 2092 (16.7) | 114 (29.1) | <0.001 |
| **Prior cerebrovascular accident, n (%)** |  | 980 (7.8) | 62 (15.8) | <0.001 |
| **Previous cardiac interventions, n (%)** |  | 3672 (29.3) | 195 (49.7) | <0.001 |
| **Cardiogenic shock, n (%)** |  | 103 (0.8) | 85 (21.7) | <0.001 |
| **Preoperative CPR, n (%)** |  | 50 (0.4) | 11 (2.8) | <0.001 |
| **NYHA classification, n (%)** | **Class I** | 8050 (64.2) | 140 (35.7) | <0.001 |
|  | **Class II** | 1795 (14.3) | 39 (9.9) |  |
|  | **Class III** | 1409 (11.2) | 75 (19.1) |  |
|  | **Class IV** | 1287 (10.3) | 138 (35.2) |  |
| **Inotropes within 48 hours, n (%)** |  | 239 (1.9) | 102 (26.0) | <0.001 |
| **Steroids within 24 hours, n (%)** |  | 324 (2.6) | 31 (7.9) | <0.001 |
| **Glycoprotein IIb/IIIa inhibitor within 24 hours, n (%)** |  | 165 (1.3) | 5 (1.3) | 0.876 |
| **ADP inhibitors within 5 days, n (%)** |  | 260 (2.1) | 22 (5.6) | <0.001 |
| **Last hematocrit, median [Q1,Q3]** |  | 39.5 [35.0,43.1] | 35.7 [31.0,40.5] | <0.001 |
| **Last white blood cell count, median [Q1,Q3]** |  | 7.3 [5.9,9.0] | 8.4 [6.4,11.1] | <0.001 |
| **Last platelet count, median [Q1,Q3]** |  | 209K [169K,257K] | 205K [150K,260K] | 0.007 |
| **Number of diseased major coronary arteries, n** | **None** | 5074 (40.5) | 149 (38.0) | 0.172 |
| (%) | **1** | 1077 (8.6) | 37 (9.4) |  |
|  | **2** | 1758 (14.0) | 44 (11.2) |  |
|  | **3** | 4632 (36.9) | 162 (41.3) |  |
| **Ejection fraction, median [Q1,Q3]** |  | 60 [53,67] | 55 [32,64] | **<0.001** |
| **Aortic stenosis, n (%)** |  | 3524 (28.1) | 125 (31.9) | 0.113 |
| **Mitral stenosis, n (%)** |  | 424 (3.4) | 29 (7.4) | **<0.001** |
| **Aortic valve insufficiency, n (%)** |  | 7490 (59.7) | 277 (70.7) | **<0.001** |
| **Mitral valve insufficiency, n (%)** |  | 10849 (86.5) | 365 (93.1) | **<0.001** |
| **Tricuspid valve insufficiency, n (%)** |  | 10549 (84.1) | 361 (92.1) | **<0.001** |
| **Preoperative status, n (%)** | **Elective** | 7426 (59.2) | 102 (26.0) | **<0.001** |
|  | **Urgent** | 4943 (39.4) | 209 (53.3) |  |
|  | **Emergent** | 170 (1.4) | 70 (17.9) |  |
|  | **Emergent Salvage** | 2 (0.0) | 11 (2.8) |  |
| **Incidence of cardiac surgery, n (%)** | **1st** | 9918 (79.1) | 260 (66.3) | **<0.001** |
|  | **1st re-op** | 1434 (11.4) | 101 (25.8) |  |
|  | **2nd or more re-op** | 1189 (9.5) | 31 (7.9) |  |
| **Cross clamp time (min), median [Q1,Q3]** |  | 88 [57,118] | 87 [0,150] | 0.848 |
| **Cardiopulmonary bypass time (min), median [Q1,Q3]** |  | 120 [89,157] | 156 [78,242] | **<0.001** |
| **CABG performed, n (%)** |  | 6620 (52.8) | 177 (45.2) | **0.003** |
| **Valve procedure performed, n (%)** |  | 5989 (47.8) | 198 (50.5) | 0.306 |

We trained a multi-task convolutional DNN, “ECGNet”, to predict operative mortality in all types of cardiac surgical procedures, using ECG voltage alone. ECGNet achieved an AUC of 0.845 ± 0.035 when prediction was performed on all types of cardiac surgical procedures (Table 2). When prediction was performed on specific types of surgical procedures, the AUCs were 0.876 ± 0.058 (coronary artery bypass graft, CABG procedures), 0.929 ± 0.110 (valve procedures), 0.772 ± 0.104 (combined CABG and valve procedures), and 0.831 ± 0.043 (other procedures).

**Table 2:**
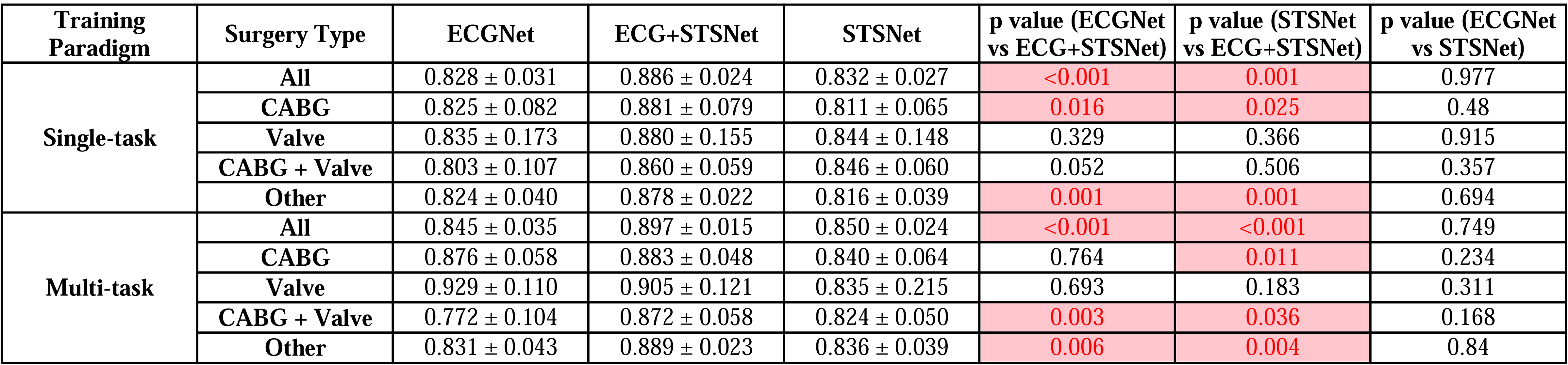
Comparison of model performances (AUC) by features used in DNNs (mortality outcomes)

| Training Paradigm | Surgery Type | ECGNet | ECG+STSNet | STSNet | p value (ECGNet vs ECG+STSNet) | p value (STSNet vs ECG+STSNet) | p value (ECGNet vs STSNet) |
| --- | --- | --- | --- | --- | --- | --- | --- |
| Single-task | All | $0.828 \pm 0.031$ | $0.886 \pm 0.024$ | $0.832 \pm 0.027$ | <0.001 | 0.001 | 0.977 |
| | CABG | $0.825 \pm 0.082$ | $0.881 \pm 0.079$ | $0.811 \pm 0.065$ | 0.016 | 0.025 | 0.48 |
| | Valve | $0.835 \pm 0.173$ | $0.880 \pm 0.155$ | $0.844 \pm 0.148$ | 0.329 | 0.366 | 0.915 |
| | CABG + Valve | $0.803 \pm 0.107$ | $0.860 \pm 0.059$ | $0.846 \pm 0.060$ | 0.052 | 0.506 | 0.357 |
| | Other | $0.824 \pm 0.040$ | $0.878 \pm 0.022$ | $0.816 \pm 0.039$ | 0.001 | 0.001 | 0.694 |
| Multi-task | All | $0.845 \pm 0.035$ | $0.897 \pm 0.015$ | $0.850 \pm 0.024$ | <0.001 | <0.001 | 0.749 |
| | CABG | $0.876 \pm 0.058$ | $0.883 \pm 0.048$ | $0.840 \pm 0.064$ | 0.764 | 0.011 | 0.234 |
| | Valve | $0.929 \pm 0.110$ | $0.905 \pm 0.121$ | $0.835 \pm 0.215$ | 0.693 | 0.183 | 0.311 |
| | CABG + Valve | $0.772 \pm 0.104$ | $0.872 \pm 0.058$ | $0.824 \pm 0.050$ | 0.003 | 0.036 | 0.168 |
| | Other | $0.831 \pm 0.043$ | $0.889 \pm 0.023$ | $0.836 \pm 0.039$ | 0.006 | 0.004 | 0.84 |

We trained a multi-task DNN, “STSNet”, to predict operative mortality in all types of cardiac surgical procedures, using STS features. STSNet achieved an AUC of 0.850 ± 0.024 when prediction was performed on all types of cardiac surgical procedures (Table 2). When prediction was performed on specific types of surgical procedures, the AUCs were 0.840 ± 0.064 (CABG procedures), 0.835 ± 0.215 (valve procedures), 0.824 ± 0.050 (combined CABG and valve procedures), and 0.836 ± 0.039 (other procedures).

Combining ECG and STS data significantly improved prediction of operative mortality compared to either data modality alone in the single-task training paradigm across all surgery types (Table 2). In the multi-task training paradigm, combining data modalities significantly improved model performance compared to either data modality alone for all, CABG, combined CABG and valve, and other procedures (Table 2). Figure 3 shows ROC, PR, and calibration curves of multi-task mortality prediction models for all surgery types.

Performant clinical prediction models are (1) discriminatory, i.e., discerns between patients who will have versus will not have an event, and (2) well-calibrated, i.e., accurately predicts absolute risk. For example, a model that is poorly calibrated may be accurate for subjects at high risk of the outcome, but less accurate for subjects at moderate or low risk. Both discrimination and calibration are important in the context of pre-surgical risk assessment (13).

To assess calibration, we report the number of expected events over the number of observed (e.g., actual) events, or E/O ratio. The E/O ratio provides the ratio of the total expected individuals to present a determined outcome over the actual number of individuals who had the specific outcome. In other words, it describes the agreement between the predicted risk and the observed risk. A perfectly calibrated model has an E/O ratio of one, which means that the model predicts the exact number of observed events. A poorly calibrated model has an E/O deviating from one, which means that the model under-predicts the risk of the outcome and vice versa (14). Evaluating the calibration of our models, we found that our models are generally well calibrated with E/O ratio close to one (Figure 3).

Multi-task convolutional DNNs or DNNs performed significantly better than single-task models when prediction of operative mortality was performed on all types of cardiac surgical procedures by STSNet and on CABG procedures by ECGNet (Supplementary Table 4). When prediction was performed on specific types of cardiac surgery, the differences in the mean AUC between the single-task and multi-task models for mortality prediction were not statistically significant in most cases. STSNet and ECGNet multi-task models can also predict secondary outcomes including postoperative stroke and renal failure within the same model (Supplementary Tables 4, 5). When predicting secondary outcomes, multi-task models performed significantly better than single-task models in certain scenarios, e.g., in the prediction of renal failure and stroke for all types of cardiac surgical procedures by ECGNet (Supplementary Table 4).

Additional model performance metrics (AUPRC, accuracy, F1 and precision) are reported in Table 3. Similar to the main reported metric of AUC (Table 2, Supplementary Tables 4, 5), combining STS and ECG data significantly improved AUPRC, accuracy, F1, and precision in the prediction of operative mortality after all procedures, as well as AUPRC, F1, and precision after CABG procedures, and AUPRC and F1 after other procedures compared to STSNet alone. Combining STS and ECG data generally significantly improved model performance in the prediction of operative mortality after all procedures (AUPRC, precision), and after other procedures (AUPRC, F1, precision) compared to ECGNet alone. The additional performance metrics of ECGNet were similar to those of STSNet after all procedures. ECGNet significantly outperformed STSNet in AUPRC, accuracy, and F1 after CABG procedures, and precision after valve procedures.

**Table 3:**
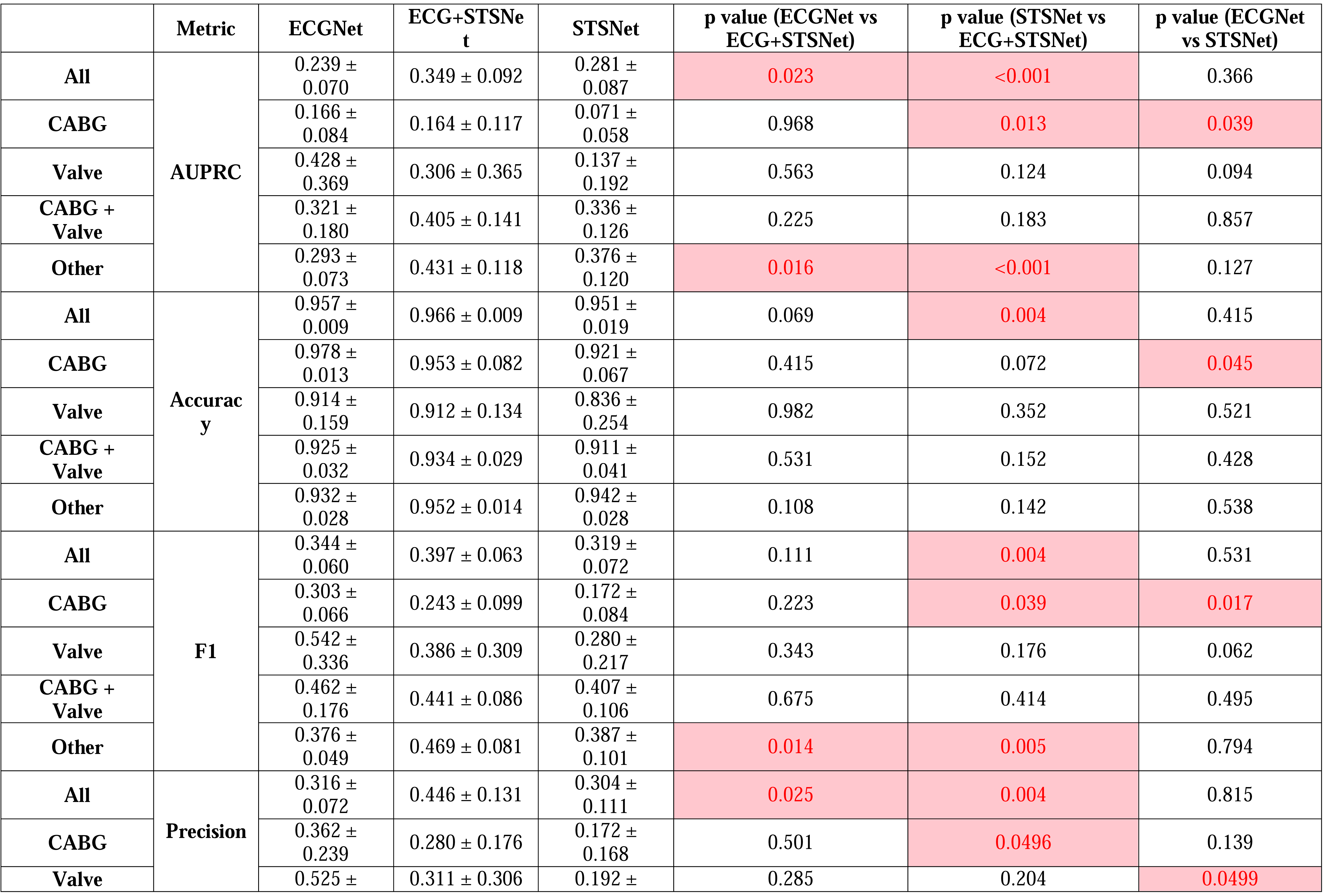
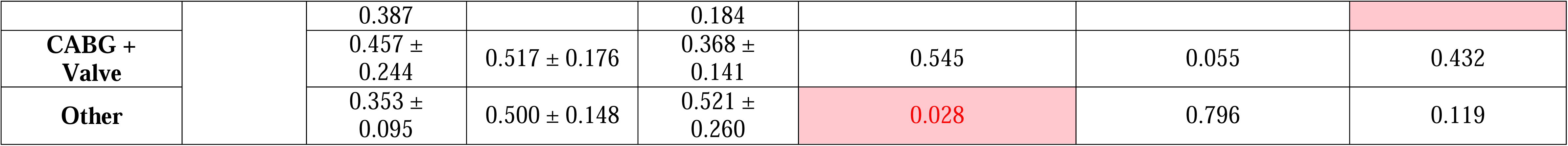
Additional model performance metrics (outcome: operative mortality)

An ablation study (Supplementary Table 6) was performed to assess how mortality prediction varied by features used in DNNs when trained in a multi-task fashion using all outcomes except one. There was no statistical significance (p-value < 0.05) on the performance when trained in a multitask fashion with all the outcomes versus removing any of them in any case except when removing stroke or long length of stay for the STSNet (denoted with an asterisk in the table). This result suggests that stroke and long length of stay might be the outcomes that contribute the most to improve operative mortality prediction.

## Discussion

The first report of training a convolutional DNN to predict all-cause 1-year mortality from ECGs was published by Raghunath et al. and included a broad cohort of patients (15). In this present study, we focus specifically on patients who underwent cardiac surgical procedures, and we investigate operative mortality and other outcomes. We hypothesized that a convolutional DNN could learn features in ECG voltage waveforms that are associated with operative mortality and other adverse outcomes. These may include presence of arrhythmias or arrhythmogenic substrate, ischemia, structural abnormalities, other impairments of cardiovascular function, or important comorbidities associated with adverse outcomes such as diabetes or obesity.

We show a convolutional DNN trained on preoperative ECGs alone - ECGNet - can predict operative mortality in a cardiac surgical population from a single institution with good performance (AUC 0.85). This result is comparable to other models using deep learning ECG data that included a larger number of inpatient procedures, including cardiac surgeries (AUC 0.85), non-cardiac surgeries, and catheterization laboratory or endoscopy suite interventions (16). Both results are higher than previously reported performance estimates of existing established risk models such as the STS score, EuroSCORE II, and ACEF score, which ranged from 0.72 to 0.80 (2, 17) and recent machine learning models using STS data, which ranged from 0.77 to 0.83 (mitral valve surgery) (18) and 0.76 to 0.78 (aortic valve surgery, up to 0.79 when concordant) (19, 20). These results support our hypothesis that the ECG voltage waveforms contain rich prognostic information related to operative mortality. For comparison, we also trained a DNN using STS features alone, STSNet, that achieved statistically similar performance to ECGNet. These combined results demonstrate the flexibility and the power of neural networks to improve peri-operative risk prediction. In situations where the patient’s ECG is available but medical history (which form a significant proportion of STS features) is limited or unavailable, e.g., in the case of an unconscious patient, ECGNet may be an alternative to STSNet, or other models using STS features, in the prediction of operative mortality.

To test the hypothesis that information in the voltage waveform could augment the predictive performance of STSNet, we trained a model using both ECG waveforms and STS features. This model performed significantly better than either STSNet or ECGNet alone, in the prediction of operative mortality in all procedures, combined CABG and valve procedures and other procedures (Table 2). The addition of other prognostically rich data modalities, such as chest radiographs or echocardiograms, may also be considered to augment the predictive performance of STSNet (21). Deep learning architectures such as convolutional neural networks enable ingestion of diverse and complementary data types in their native formats. Harnessing a fuller set of information compared to features that are manually extracted using image or signal processing techniques could further enhance risk prediction and phenotyping.

We experimented with adding summary metadata to ECGNet computed by automated ECG systems (ventricular rate, PR interval, QRS duration, QT interval, P axis, R axis, and T axis). However, adding ECG metadata did not significantly improve model performance of ECGNet (data not shown), suggesting the high-dimensional latent representation of the ECG voltage that is learned by ECGNet is redundant with the information in computed ECG metadata.

We compared the performance of multi-task and single-task neural networks, in the context of cardiac surgical procedures. Existing conventional risk models had been created in a single-task fashion. We note the AUCs of multi-task models were comparable or significantly higher than single-task models, and there were no cases where the AUCs of single-task models were significantly higher than multi-task models. Multitask learning assumes some relation between the various tasks (i.e., clinical outcomes to predict). Empirically and theoretically, jointly learning multiple tasks can improve performance versus learning each task independently by leveraging hidden information between phenotypes, outcomes, and patient states that are not explicitly described or modeled (22-24). This study demonstrates the ability of deep learning to capture complex relationships, while eliminating the need to train individual models for each outcome.

For patients undergoing non-major procedure types, STS risk scores were not available and hence not directly comparable against our models. Moreover, we were not able to obtain missing scores from past versions of the STS risk calculator due to changing data specifications per version. This latter source of missingness precludes the comparison of ECGNet directly with the STS risk score within subcohorts of specific procedure types. Comparing ECGNet with published performance metrics of the STS risk score within subcohorts required separate ECGNet models trained on subcohorts for a fair comparison, but stratifying by specific procedure type within our cohort resulted in sample sizes that were too small to fit performant and generalizable deep learning models (results not shown). A potential extension of this work may be to pretrain ECGNet using all patients and subsequently fine-tune the model on patient subcohorts; however, this procedure involves additional challenges which we have not yet investigated. Therefore, to compare ECGNet with a baseline risk score, we trained a DNN on the features used in the STS risk score, STSNet.

This STSNet model trained on STS features alone achieved an AUC of 0.85 and can be applied to any cardiac surgical procedure, including those procedures that do not currently have STS risk calculators. To our knowledge the performance of this model surpasses all prior benchmarks of discriminatory performance for mortality using these features, even when models are trained on specific surgical case types (2, 17, 25). The simplicity of having a single model that can be applied for any surgical case type has significant appeal, particularly when modern cardiac surgical procedures are becoming more complex and diverse. We previously demonstrated models that can be applied to cardiac surgical cases that do not have conventional STS risk models (25). The current STSNet using a DNN approach exceeds performance of these models.

While STSNet performed well and demonstrates potential for a deep learning method to further improve upon the existing STS risk calculator, this model was trained using data from only one quaternary academic institution with a unique case mix with limited population size. Evaluating the ability to generalize to other institutions with different case mixes, as well as the performance on well-defined patient sub-cohorts, are important next steps before broader dissemination and use.

Only 394 (3%) of 12,933 patients in our dataset experienced operative mortality. A classifier trained on class imbalanced data in which the prevalence of the primary outcome is small can favor the majority class, i.e., the more prevalent outcome (26). Bias in the trained classifier can result in poor discrimination and/or calibration on the classification task of interest. Strategies have been proposed to address class imbalance, such as undersampling or underweighing the majority class, doing the opposite for the minority class, and generative methods which seek to shift the distribution of training data in a manner that reduces classifier bias (27, 28). Interestingly, despite the low prevalence of the primary outcome in our data, applying a well-established method called Synthetic Minority Oversampling Technique (SMOTE) to address class imbalance did not improve discriminatory performance of our classifier (25). Moreover, models trained on ECG waveforms achieved both good discrimination and calibration performance, notably even at lower expected event probabilities (Figure 3). Nonetheless, the clinical importance of post-operative mortality is significant and further exploration of methods to improve performance of classifying or predicting low prevalence outcomes is warranted given their significant clinical consequences in cardiac surgical care. One alternative approach utilized in deep learning research is to inversely weigh the loss with the prevalence of the label: if there are N total samples and R have an outcome of postoperative mortality, the loss for the samples with this outcome is weighed by N/R and the loss for the samples without this outcome is weighted by N / (N-R) (29).

This study faced several limitations. Although our ECG dataset is relatively large at 4.7 million distinct studies, the intersection between these data and patients in our STS database for whom we have adjudicated outcomes is substantially smaller at 12,933 patients. A potential source of missingness for preoperative ECGs is the situation where a patient had a preoperative ECG outside of our hospital and was referred or transferred for surgery. These patients’ ECGs would not be available in our electronic dataset but would have likely been available in some other viewing format to the clinicians at the time of surgery. A larger sample size may have increased confidence that these results generalize to a broader population. It also would have increased the probability of rarer phenotypes in the training set and could potentially boost the performance of ECGNet, especially when training models on sub-cohorts. Most reports in the literature that utilize DNNs for the analysis of ECGs have sample sizes in the hundreds of thousands, and some in the millions. We used all available data for training, validation, and test sets and did not assess how performance varied with decreasing sample size. However, it is plausible for performance to increase with a larger sample size or more sophisticated data augmentation methods.

Another limitation of this general approach is the need to digitize ECGs, modifying the format to be suitable for computational approaches such as deep learning. Although most major academic medical centers maintain STS databases with tabular features such as those used in this work, digitization of the accompanying ECGs may pose substantial logistical and financial barriers. Nonetheless, we believe that quantitative analysis of native resolution digital data will become an increasingly prevalent component of research and patient care in the future.

Selecting a time window between the preoperative ECG and the cardiac surgical procedure involves a tradeoff between sample size and phenotypic precision. We trained a model on ECGs from the 60-day preoperative window. A larger window increases the number of ECGs with which to train a model, but also increases the probability that an ECG is far in time from the cardiac surgical procedure, and thus less representative of the patient’s cardiovascular phenotype proximal to when a prediction of mortality is most useful.

The prediction of secondary outcomes is variable, at times single-task models performed better than multi-task models for reoperation, and long length of stay. It may be possible to weigh the loss function against outcomes in a different, non-equal way and/or modify early stopping parameters, to improve performance on non-mortality outcomes. It may also be possible to incorporate attention and transformer layers in our models, which have been shown promise in recent studies (30, 31).

## Conclusions

We trained ECGNet, a convolutional DNN, to predict the risk of operative mortality and other adverse outcomes after cardiac surgical procedures using 30,877 preoperative ECGs (native resolution digital waveform data) from 12,933 patients. ECGNet achieved an overall AUC of 0.85 for any procedure and up to an AUC of 0.93 for valve procedures (Central Illustration). We also report excellent predictive performance of a DNN model trained using STS features (AUC 0.85), demonstrating the potential for deep learning in predicting postoperative outcomes. When STSNet and ECGNet were combined, the AUC for all procedures was 0.90, which is significantly higher than either ECGNet or STSNet alone and demonstrates the potential for improved risk prediction using multidimensional data sources and deep neural networks. Both ECGNet and STSNet are able to predict multiple outcomes using the same model at the same time and are not limited to certain types of cardiac surgical procedures.

## Abbreviations

ECG: Electrocardiogram
STS: Society for Thoracic Surgeons
AUC: Area under the receiver operating characteristic curve
CABG: Coronary artery bypass graft
DNN: Deep neural network
MGH: Massachusetts General Hospital
ROC: Receiver operating characteristic
E/O: “Expected to observed

## Data availability

The datasets generated and/or analyzed during the current study are not publicly available due to the Health Insurance Portability and Accountability Act of 1996 (HIPAA) Privacy Rule protecting individually identifiable health information but are available from the corresponding authors on reasonable request with appropriate data sharing agreements in place.

## Code availability

The underlying code for this study will be made available in GitHub after publication and can be accessed via this link https://github.com/aguirre-lab/ml4c3.

## Acknowledgements

A.D.A. discloses support for the research of this work from Controlled Risk Insurance Company / Risk Management Foundation (CRICO) and the Massachusetts General Hospital Corrigan Minehan Heart Center Hassenfeld Cardiovascular Scholar Award. Images in the Central Illustration are used with license from Adobe Stock, Shutterstock and Flaticon.

## Author contributions

CSO, ER, TS, CMS, and ADA designed the study. CSO, ER, RPV, and ADA analyzed and interpreted the data. CSO, ER, and ADA drafted the article. All authors read, critically reviewed, and approved the final manuscript.

## Figure Legends

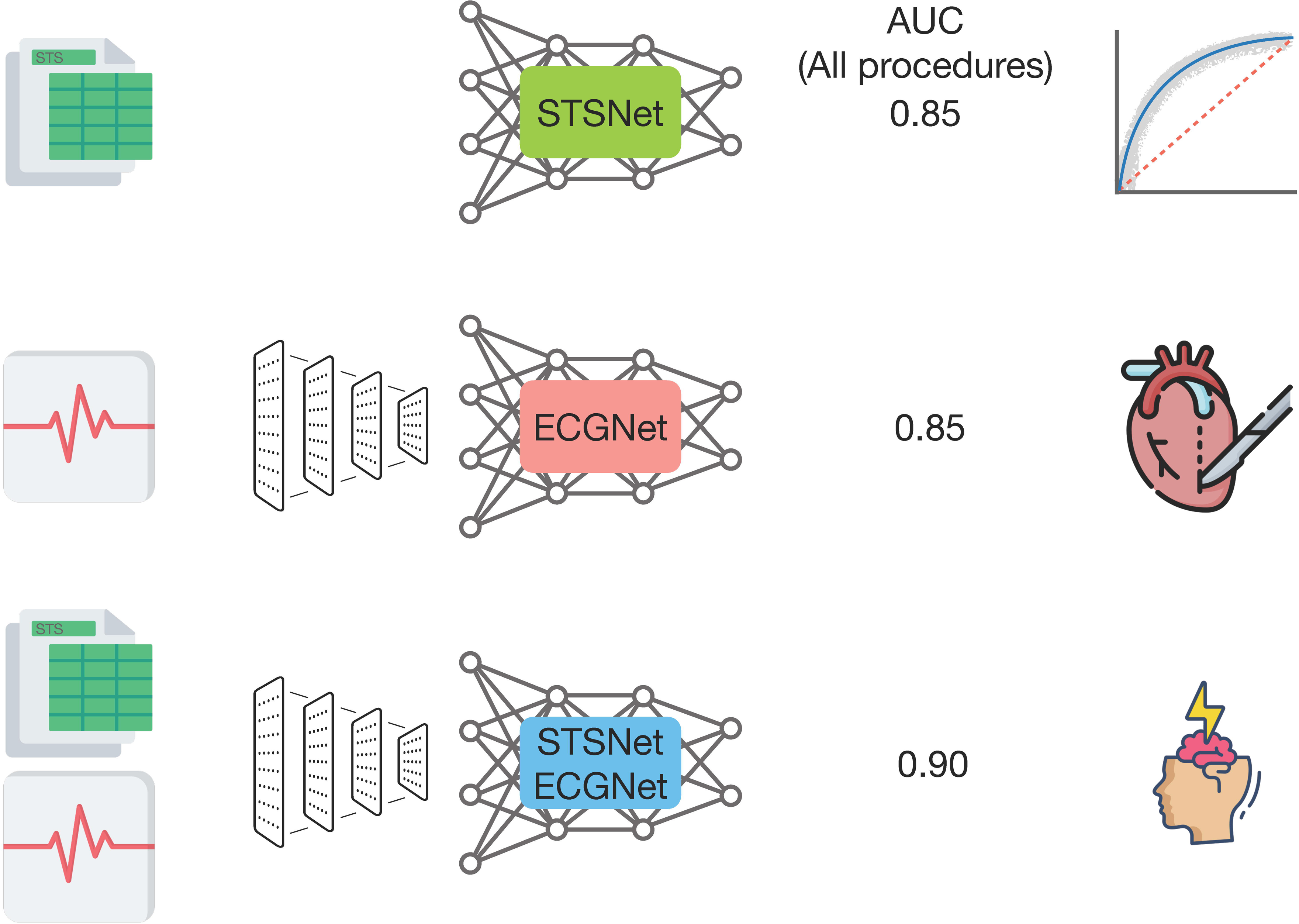
Central Illustration: We trained ECGNet and STSNet, deep neural networks that predict the risk of operative mortality and other adverse outcomes after cardiac surgical procedures. Both ECGNet and STSNet are not limited to certain types of cardiac surgical procedures and can predict multiple outcomes using the same model at the same time. Combining ECGNet and STSNet achieved a mean AUC of 0.90 for predicting operative mortality after all procedures, which is significantly higher than either ECGNet or STSNet alone.

**Supplementary Figure 1:**
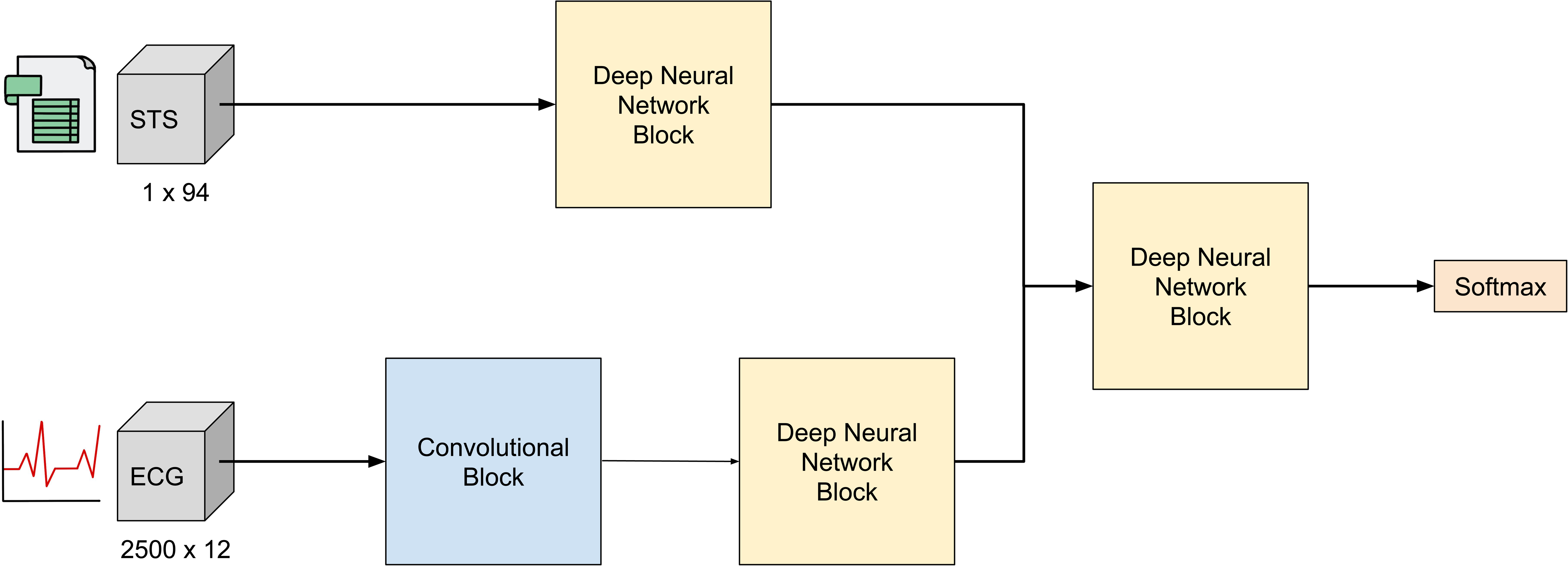
Base convolutional neural network architecture to combine ECG and STS features. We optimized a convolutional neural network composed by one convolutional block plus several deep neural network blocks. The convolutional block was composed by a convolutional layer with 32 filters and multiple dense blocks of which parameters were optimized. The deep neural network blocks are tuned independently and can contain multiple dense layers or can be ignored as described in the hyperparameter optimization section.

