## Supplementary Information for "Predicting adverse outcomes after cardiac surgery using multi-task deep neural networks, clinical features, and electrocardiograms"

**Title**:

Assistant Professor of Surgery

Division of Surgical Outcomes

Surgery Center for Health Services and Outcomes Research

Department of Surgery, Yale School of Medicine

100 Church Street South

Suite F208

New Haven, CT 06510

**Supplementary Information**

Supplementary Methods

Supplementary Tables 1 - 6

Supplementary Figure 1

**Supplementary Methods**

*Hyperparameter optimization*

For MLP architectures (e.g. models that used tabular features but did not use voltage as input), the following parameters and choices were randomly searched: optimizer (stochastic gradient descent, Adam, RAdam, AdamW), activation function (ReLU, swish), learning rate (0.0001, 0.0003, 0.0005, 0.001), dropout rate in dense layers (0.1, 0.2, 0.3, 0.4, 0.5, 0.6), and dense layer count and number of neurons ([10, 10], [100, 100], [64, 32], [64, 16], [16, 64], [64, 32, 16]). For convolutional neural network architectures (e.g. models that used voltage as input), the following parameters and choices were randomly searched: optimizer (stochastic gradient descent, Adam, RAdam, AdamW), activation function (ReLU, swish), learning rate (0.0001, 0.0003, 0.0005, 0.001), dropout rate in dense layers (0.1, 0.2, 0.3, 0.4, 0.5, 0.6), dense layer count and number of neurons (no dense layers, [64, 32], [64, 16], [32, 16], [64, 32, 16]), 1D convolutional filter size (30, 50, 70, 100), spatial dropout rate (0.15, 0.2, 0.25, 0.3), number of dense convolutional blocks and the number of filters per dense convolutional block ([12, 24, 48, 96, 192], [12, 24, 48, 96]), number of layers within each dense convolutional block (2, 3), and convolutional pooling factor (2, 3, 4).

*Feature Selection*

The list of features used in this study was derived from a list of features found to be predictive in parsimonious models (after feature selection and dimension reduction) in our previously published work (25). The full variable list used in our previously published work (25) was originally selected from a list of candidate predictors selected by a surgeon panel (1, 2) and underwent backward selection and optimal coding (1, 2).

*ECG and tabular data combination*

For architectures combining ECG and STS features, different base architectures have been tried to test the possible ways to combine tabular and time-series features. To this end, we have optimized the parameters of a network with a convolutional block plus a deep neural network block to process the ECG time series, a deep neural network block to process the tabular features, and a last deep neural network block that processes the combination of the outputs of each one of the deep neural blocks (Supplementary Figure 1). The parameters of each deep neural block were tuned independently. The parameters and choices are shown in the hyperparameter optimization section. The model architecture that was finally selected is detailed in Figure 2.

**Supplementary Table 1: Features used to train STSNet**

| **Feature** | **Variable type** |
| --- | --- |
| ADP inhibitors within past 5 days | Binary |
| Age | Continuous |
| Aortic stenosis | Binary |
| Aortic valve insufficiency or regurgitation | Binary |
| Cardiogenic shock | Binary |
| Cerebrovascular disease | Binary |
| Chronic lung disease | Binary |
| Diabetes | Binary |
| Dialysis | Binary |
| Ejection fraction | Continuous |
| Ethnicity | Binary |
| Gender (Male) | Binary |
| Glycoprotein IIb/IIIa inhibitor within past 24 hours | Binary |
| Heart failure | Binary |
| Height (cm) | Continuous |
| Hypertension | Binary |
| Immunocompromised status | Binary |
| Incidence of cardiac surgery | Binary |
| Infective endocarditis type | Binary |
| Inotropes within 48 hours | Binary |
| Last creatinine level | Continuous |
| Last hematocrit | Continuous |
| Last platelet count | Continuous |
| Last WBC count | Continuous |
| Mitral stenosis | Binary |
| Mitral valve insufficiency or regurgitation | Binary |
| More than 6 hours between previous PCI and current procedure | Binary |
| Number of diseased vessels | Categorical |
| NYHA classification | Categorical |
| Peripheral arterial disease | Binary |
| Preop CPR | Binary |
| Preop status | Binary |
| Previous CABG | Binary |
| Previous cardiac interventions | Binary |
| Previous PCI | Binary |
| Previous transient ischemic attack | Binary |
| Previous valve procedure | Binary |
| Prior carotid surgery | Binary |
| Prior cerebrovascular accident | Binary |
| Race (Asian) | Binary |
| Race (Black) | Binary |
| Race (Caucasian) | Binary |
| Race (Native American) | Binary |
| Race (Other or Native Pacific) | Binary |
| Steroids within past 24 hours | Binary |
| Tricuspid valve insufficiency or regurgitation | Binary |
| Weight (kg) | Continuous |

**Supplementary Table 2: Optimal deep neural network architectures determined via random hyperparameter search**

| **Parameter** | **ECGNet** | **STSNet** | **ECG+STSNet** |
| --- | --- | --- | --- |
| **Filter size** | 30 |  | 30 |
| **Number of filters per convolutional block** | 32 |  | 32 |
| **Convolutional block size** | 1 |  | 1 |
| **Number of filters per dense convolutional blocks** | 12, 24, 48, 96 |  | 12, 24, 48, 96 |
| **Dense convolutional block size** | 2 |  | 2 |
| **Spatial dropout rate** | 0.25 |  | 0.25 |
| **Pooling after convolutional block** | max pool 3 |  | max pool 3 |
| **Number of neurons per fully connected layer** | 64, 32 | 64, 32, 16 | 64, 32 |
| **Dropout rate** | 0.25 | 0.2 | 0.25 |
| **Activation** | relu | relu | relu |
| **Optimizer** | adamw | adam | adamw |
| **Learning rate** | 0.0001 | 0.0001 | 0.0001 |
| **Weight decay** | 0.0001 |  | 0.0001 |
| **L2 regularization** | 0.0001 |  | 0.0001 |

**Supplementary Table 3: Outcomes of interest for ECGNet, STSNet and ECG+STSNet**

| **Outcome name** | **Variable type** |
| --- | --- |
| Operative mortality | Binary |
| Stroke | Binary |
| Renal failure | Binary |
| Prolonged ventilation | Binary |
| Reoperation | Binary |
| Long length of stay | Binary |

**Supplementary Table 4: Comparison of model performances (AUC), single-task DNNs vs multi-task DNNs (All outcomes)**

|  |  | **ECGNet** | | | **ECG+STSNet** | | | **STSNet** | | |
| --- | --- | --- | --- | --- | --- | --- | --- | --- | --- | --- |
|  |  | **Multi-task** | **Single-task** | **p value (Multi-task vs Single-task)** | **Multi-task** | **Single-task** | **p value (Multi-task vs Single-task)** | **Multi-task** | **Single-task** | **p value (Multi-task vs Single-task)** |
| **Operative mortality** | **All** | 0.845 ± 0.035 | 0.828 ± 0.031 | 0.200 | 0.897 ± 0.015 | 0.886 ± 0.024 | 0.193 | 0.850 ± 0.024 | 0.832 ± 0.027 | 0.013 |
|  | **CABG** | 0.876 ± 0.058 | 0.825 ± 0.082 | 0.012 | 0.883 ± 0.048 | 0.881 ± 0.079 | 0.949 | 0.840 ± 0.064 | 0.811 ± 0.065 | 0.079 |
|  | **Valve** | 0.929 ± 0.110 | 0.835 ± 0.173 | 0.235 | 0.905 ± 0.121 | 0.880 ± 0.155 | 0.655 | 0.835 ± 0.215 | 0.844 ± 0.148 | 0.778 |
|  | **CABG + Valve** | 0.772 ± 0.104 | 0.803 ± 0.107 | 0.098 | 0.872 ± 0.058 | 0.860 ± 0.059 | 0.474 | 0.824 ± 0.050 | 0.846 ± 0.060 | 0.170 |
|  | **Other** | 0.831 ± 0.043 | 0.824 ± 0.040 | 0.541 | 0.889 ± 0.023 | 0.878 ± 0.022 | 0.18 | 0.836 ± 0.039 | 0.816 ± 0.039 | 0.052 |
| **Stroke** | **All** | 0.602 ± 0.034 | 0.545 ± 0.038 | 0.003 | 0.647 ± 0.033 | 0.566 ± 0.042 | 0.006 | 0.642 ± 0.041 | 0.603 ± 0.059 | 0.143 |
|  | **CABG** | 0.548 ± 0.073 | 0.614 ± 0.052 | 0.071 | 0.609 ± 0.131 | 0.541 ± 0.072 | 0.189 | 0.574 ± 0.132 | 0.557 ± 0.084 | 0.769 |
|  | **Valve** | 0.638 ± 0.136 | 0.545 ± 0.116 | 0.138 | 0.755 ± 0.138 | 0.500 ± 0.107 | 0.001 | 0.722 ± 0.179 | 0.697 ± 0.123 | 0.744 |
|  | **CABG + Valve** | 0.528 ± 0.091 | 0.477 ± 0.172 | 0.502 | 0.520 ± 0.102 | 0.561 ± 0.087 | 0.363 | 0.553 ± 0.122 | 0.558 ± 0.142 | 0.95 |
|  | **Other** | 0.603 ± 0.070 | 0.538 ± 0.065 | 0.016 | 0.623 ± 0.066 | 0.567 ± 0.069 | 0.133 | 0.640 ± 0.062 | 0.584 ± 0.088 | 0.138 |
| **Renal failure** | **All** | 0.751 ± 0.024 | 0.716 ± 0.048 | 0.032 | 0.809 ± 0.021 | 0.809 ± 0.032 | 0.973 | 0.769 ± 0.027 | 0.759 ± 0.057 | 0.713 |
|  | **CABG** | 0.776 ± 0.067 | 0.722 ± 0.089 | 0.192 | 0.819 ± 0.051 | 0.822 ± 0.055 | 0.91 | 0.747 ± 0.050 | 0.740 ± 0.093 | 0.873 |
|  | **Valve** | 0.731 ± 0.132 | 0.769 ± 0.111 | 0.679 | 0.796 ± 0.211 | 0.865 ± 0.096 | 0.406 | 0.790 ± 0.121 | 0.746 ± 0.135 | 0.528 |
|  | **CABG + Valve** | 0.745 ± 0.115 | 0.704 ± 0.110 | 0.226 | 0.796 ± 0.062 | 0.788 ± 0.054 | 0.729 | 0.745 ± 0.061 | 0.761 ± 0.075 | 0.607 |
|  | **Other** | 0.724 ± 0.040 | 0.688 ± 0.069 | 0.161 | 0.784 ± 0.019 | 0.782 ± 0.053 | 0.925 | 0.762 ± 0.031 | 0.748 ± 0.062 | 0.563 |
| **Long length of stay** | **All** | 0.688 ± 0.028 | 0.694 ± 0.028 | 0.627 | 0.777 ± 0.021 | 0.792 ± 0.019 | 0.11 | 0.761 ± 0.014 | 0.785 ± 0.023 | 0.048 |
|  | **CABG** | 0.693 ± 0.023 | 0.707 ± 0.063 | 0.522 | 0.750 ± 0.023 | 0.775 ± 0.052 | 0.183 | 0.704 ± 0.033 | 0.772 ± 0.047 | 0.005 |
|  | **Valve** | 0.697 ± 0.079 | 0.708 ± 0.065 | 0.715 | 0.810 ± 0.052 | 0.816 ± 0.063 | 0.803 | 0.822 ± 0.041 | 0.808 ± 0.075 | 0.598 |
|  | **CABG + Valve** | 0.622 ± 0.077 | 0.670 ± 0.059 | 0.057 | 0.729 ± 0.024 | 0.781 ± 0.049 | 0.008 | 0.741 ± 0.030 | 0.758 ± 0.056 | 0.417 |
|  | **Other** | 0.680 ± 0.032 | 0.662 ± 0.041 | 0.404 | 0.774 ± 0.028 | 0.781 ± 0.030 | 0.676 | 0.757 ± 0.020 | 0.774 ± 0.034 | 0.309 |
| **Reoperation** | **All** | 0.620 ± 0.014 | 0.633 ± 0.011 | 0.135 | 0.727 ± 0.013 | 0.738 ± 0.009 | 0.103 | 0.712 ± 0.013 | 0.724 ± 0.013 | 0.104 |
|  | **CABG** | 0.608 ± 0.032 | 0.615 ± 0.015 | 0.649 | 0.734 ± 0.021 | 0.753 ± 0.018 | 0.126 | 0.721 ± 0.017 | 0.747 ± 0.020 | 0.046 |
|  | **Valve** | 0.649 ± 0.046 | 0.651 ± 0.044 | 0.965 | 0.707 ± 0.029 | 0.706 ± 0.038 | 0.938 | 0.668 ± 0.025 | 0.665 ± 0.028 | 0.868 |
|  | **CABG + Valve** | 0.601 ± 0.056 | 0.588 ± 0.033 | 0.65 | 0.656 ± 0.039 | 0.687 ± 0.025 | 0.087 | 0.637 ± 0.049 | 0.671 ± 0.031 | 0.117 |
|  | **Other** | 0.615 ± 0.021 | 0.644 ± 0.022 | 0.006 | 0.735 ± 0.024 | 0.740 ± 0.013 | 0.544 | 0.726 ± 0.024 | 0.727 ± 0.018 | 0.931 |
| **Prolonged ventilation** | **All** | 0.706 ± 0.037 | 0.685 ± 0.024 | 0.193 | 0.787 ± 0.025 | 0.771 ± 0.027 | 0.209 | 0.772 ± 0.024 | 0.766 ± 0.021 | 0.671 |
|  | **CABG** | 0.723 ± 0.039 | 0.691 ± 0.066 | 0.189 | 0.762 ± 0.025 | 0.746 ± 0.042 | 0.409 | 0.736 ± 0.024 | 0.743 ± 0.033 | 0.68 |
|  | **Valve** | 0.649 ± 0.102 | 0.670 ± 0.046 | 0.628 | 0.781 ± 0.057 | 0.779 ± 0.076 | 0.955 | 0.763 ± 0.053 | 0.772 ± 0.089 | 0.842 |
|  | **CABG + Valve** | 0.675 ± 0.075 | 0.650 ± 0.047 | 0.374 | 0.761 ± 0.047 | 0.730 ± 0.036 | 0.023 | 0.748 ± 0.052 | 0.718 ± 0.032 | 0.074 |
|  | **Other** | 0.703 ± 0.047 | 0.667 ± 0.013 | 0.069 | 0.797 ± 0.035 | 0.779 ± 0.036 | 0.252 | 0.784 ± 0.025 | 0.780 ± 0.029 | 0.809 |

**Supplementary Table 5: Comparison of model performances (AUC) by features used in DNNs (Non-mortality outcomes)**

|  |  | **Single-task** | | | | | | **Multi-task** | | | | | |
| --- | --- | --- | --- | --- | --- | --- | --- | --- | --- | --- | --- | --- | --- |
| **Outcome** | **Surgery Type** | **ECGNet** | **ECG+STSNet** | **STSNet** | **p value (ECGNet vs ECG+STSNet)** | **p value (STSNet vs ECG+STSNet)** | **p value (ECGNet vs STSNet)** | **ECGNet** | **ECG+STSNet** | **STSNet** | **p value (ECGNet vs ECG+STSNet)** | **p value (STSNet vs ECG+STSNet)** | **p value (ECGNet vs STSNet)** |
| **Stroke** | **All** | 0.545 ± 0.038 | 0.566 ± 0.042 | 0.603 ± 0.059 | 0.306 | 0.161 | 0.011 | 0.602 ± 0.034 | 0.647 ± 0.033 | 0.642 ± 0.041 | <0.001 | 0.504 | 0.002 |
|  | **CABG** | 0.614 ± 0.052 | 0.541 ± 0.072 | 0.557 ± 0.084 | 0.011 | 0.709 | 0.137 | 0.548 ± 0.073 | 0.609 ± 0.131 | 0.574 ± 0.132 | 0.028 | 0.187 | 0.417 |
|  | **Valve** | 0.545 ± 0.116 | 0.500 ± 0.107 | 0.697 ± 0.123 | 0.45 | 0.005 | 0.033 | 0.638 ± 0.136 | 0.755 ± 0.138 | 0.722 ± 0.179 | 0.012 | 0.318 | 0.127 |
|  | **CABG + Valve** | 0.477 ± 0.172 | 0.561 ± 0.087 | 0.558 ± 0.142 | 0.148 | 0.962 | 0.378 | 0.528 ± 0.091 | 0.520 ± 0.102 | 0.553 ± 0.122 | 0.847 | 0.302 | 0.655 |
|  | **Other** | 0.538 ± 0.065 | 0.567 ± 0.069 | 0.584 ± 0.088 | 0.338 | 0.595 | 0.135 | 0.603 ± 0.070 | 0.623 ± 0.066 | 0.640 ± 0.062 | 0.074 | 0.293 | 0.066 |
| **Renal failure** | **All** | 0.716 ± 0.048 | 0.809 ± 0.032 | 0.759 ± 0.057 | <0.001 | 0.037 | 0.077 | 0.751 ± 0.024 | 0.809 ± 0.021 | 0.769 ± 0.027 | <0.001 | <0.001 | 0.26 |
|  | **CABG** | 0.722 ± 0.089 | 0.822 ± 0.055 | 0.740 ± 0.093 | 0.02 | 0.032 | 0.62 | 0.776 ± 0.067 | 0.819 ± 0.051 | 0.747 ± 0.050 | 0.059 | 0.003 | 0.342 |
|  | **Valve** | 0.769 ± 0.111 | 0.865 ± 0.096 | 0.746 ± 0.135 | 0.069 | 0.052 | 0.723 | 0.731 ± 0.132 | 0.796 ± 0.211 | 0.790 ± 0.121 | 0.315 | 0.915 | 0.33 |
|  | **CABG + Valve** | 0.704 ± 0.110 | 0.788 ± 0.054 | 0.761 ± 0.075 | 0.012 | 0.296 | 0.215 | 0.745 ± 0.115 | 0.796 ± 0.062 | 0.745 ± 0.061 | 0.055 | 0.011 | 0.999 |
|  | **Other** | 0.688 ± 0.069 | 0.782 ± 0.053 | 0.748 ± 0.062 | <0.001 | 0.168 | 0.079 | 0.724 ± 0.040 | 0.784 ± 0.019 | 0.762 ± 0.031 | 0.001 | 0.049 | 0.061 |
| **Long length of stay** | **All** | 0.694 ± 0.028 | 0.792 ± 0.019 | 0.785 ± 0.023 | <0.001 | 0.168 | <0.001 | 0.688 ± 0.028 | 0.777 ± 0.021 | 0.761 ± 0.014 | <0.001 | 0.019 | <0.001 |
|  | **CABG** | 0.707 ± 0.063 | 0.775 ± 0.052 | 0.772 ± 0.047 | 0.002 | 0.819 | 0.012 | 0.693 ± 0.023 | 0.750 ± 0.023 | 0.704 ± 0.033 | <0.001 | <0.001 | 0.507 |
|  | **Valve** | 0.708 ± 0.065 | 0.816 ± 0.063 | 0.808 ± 0.075 | 0.003 | 0.459 | 0.006 | 0.697 ± 0.079 | 0.810 ± 0.052 | 0.822 ± 0.041 | 0.001 | 0.268 | 0.001 |
|  | **CABG + Valve** | 0.670 ± 0.059 | 0.781 ± 0.049 | 0.758 ± 0.056 | <0.001 | 0.071 | 0.008 | 0.622 ± 0.077 | 0.729 ± 0.024 | 0.741 ± 0.030 | 0.002 | 0.229 | 0.006 |
|  | **Other** | 0.662 ± 0.041 | 0.781 ± 0.030 | 0.774 ± 0.034 | <0.001 | 0.443 | <0.001 | 0.680 ± 0.032 | 0.774 ± 0.028 | 0.757 ± 0.020 | <0.001 | 0.066 | <0.001 |
| **Reoperation** | **All** | 0.633 ± 0.011 | 0.738 ± 0.009 | 0.724 ± 0.013 | <0.001 | 0.002 | <0.001 | 0.620 ± 0.014 | 0.727 ± 0.013 | 0.712 ± 0.013 | <0.001 | 0.001 | <0.001 |
|  | **CABG** | 0.615 ± 0.015 | 0.753 ± 0.018 | 0.747 ± 0.020 | <0.001 | 0.169 | <0.001 | 0.608 ± 0.032 | 0.734 ± 0.021 | 0.721 ± 0.017 | <0.001 | 0.004 | <0.001 |
|  | **Valve** | 0.651 ± 0.044 | 0.706 ± 0.038 | 0.665 ± 0.028 | 0.01 | 0.002 | 0.35 | 0.649 ± 0.046 | 0.707 ± 0.029 | 0.668 ± 0.025 | <0.001 | <0.001 | 0.093 |
|  | **CABG + Valve** | 0.588 ± 0.033 | 0.687 ± 0.025 | 0.671 ± 0.031 | 0.001 | 0.043 | 0.003 | 0.601 ± 0.056 | 0.656 ± 0.039 | 0.637 ± 0.049 | 0.004 | 0.084 | 0.11 |
|  | **Other** | 0.644 ± 0.022 | 0.740 ± 0.013 | 0.727 ± 0.018 | <0.001 | 0.007 | <0.001 | 0.615 ± 0.021 | 0.735 ± 0.024 | 0.726 ± 0.024 | <0.001 | 0.063 | <0.001 |
| **Prolonged ventilation** | **All** | 0.685 ± 0.024 | 0.771 ± 0.027 | 0.766 ± 0.021 | <0.001 | 0.366 | <0.001 | 0.706 ± 0.037 | 0.787 ± 0.025 | 0.772 ± 0.024 | <0.001 | 0.016 | <0.001 |
|  | **CABG** | 0.691 ± 0.066 | 0.746 ± 0.042 | 0.743 ± 0.033 | 0.01 | 0.747 | 0.037 | 0.723 ± 0.039 | 0.762 ± 0.025 | 0.736 ± 0.024 | 0.009 | 0.003 | 0.325 |
|  | **Valve** | 0.670 ± 0.046 | 0.779 ± 0.076 | 0.772 ± 0.089 | 0.01 | 0.577 | 0.026 | 0.649 ± 0.102 | 0.781 ± 0.057 | 0.763 ± 0.053 | 0.002 | 0.262 | 0.013 |
|  | **CABG + Valve** | 0.650 ± 0.047 | 0.730 ± 0.036 | 0.718 ± 0.032 | <0.001 | 0.18 | 0.001 | 0.675 ± 0.075 | 0.761 ± 0.047 | 0.748 ± 0.052 | 0.004 | 0.311 | 0.052 |
|  | **Other** | 0.667 ± 0.013 | 0.779 ± 0.036 | 0.780 ± 0.029 | <0.001 | 0.741 | <0.001 | 0.703 ± 0.047 | 0.797 ± 0.035 | 0.784 ± 0.025 | <0.001 | 0.201 | <0.001 |

**Supplementary Table 6: Ablation Study. Comparison of model performances (AUC) predicting mortality by features used in DNNs when trained in a multi-task fashion using all outcomes except one. *Denotes statistical significance (p-value < 0.05) when comparing performance of the model with the corresponding baseline model.**

| **Outcomes** | **ECGNet** | **ECG+STSNet** | **STSNet** |
| --- | --- | --- | --- |
| **All (baseline)** | 0.845 ± 0.035 | 0.897 ± 0.015 | 0.850 ± 0.024 |
| **All except prolonged ventilation** | 0.841 ± 0.037 | 0.892 ± 0.015 | 0.841 ± 0.024 |
| **All except stroke** | 0.842 ± 0.035 | 0.899 ± 0.015 | 0.836 ± 0.020* |
| **All except renal failure** | 0.840 ± 0.038 | 0.898 ± 0.015 | 0.842 ± 0.016 |
| **All except long length of stay** | 0.843 ± 0.030 | 0.894 ± 0.021 | 0.839 ± 0.021* |
| **All except reoperation** | 0.852 ± 0.029 | 0.901 ± 0.014 | 0.843 ± 0.025 |
